# Polygenic overlap of physical activity with schizophrenia and Parkinson’s disease

**DOI:** 10.64898/2026.06.23.26356304

**Authors:** Kristien van der Walt, Alexey Shadrin, Markos Tesfaye, Dennis van der Meer, Ole A. Andreassen, Jaroslav Rokicki, Megan Campbell

## Abstract

Physical activity levels are increasingly investigated as candidate digital biomarkers across a range of psychiatric and neurological disorders. Physical activity is heritable, with associated genetic variants implicating genes expressed in the central nervous system. However, its shared genetic architecture with disorders such as schizophrenia and Parkinson’s disease (PD) remains poorly understood. Here, we applied complementary genome-wide methods to investigate the genetic overlap between physical activity and these disorders. We used a genome-wide association study (GWAS) of objectively measured physical activity (n = 89,683), enhanced through multi-trait GWAS incorporating self-reported activity measures (n = 124,842–377,234). In cross-trait analysis with recent GWAS of schizophrenia (n = 53,386 cases, 77,258 controls) and PD (63,555 cases, 17,700 proxy cases, and 1,746,386 controls), we find that while genetic correlations are negligible (rg=-0.02–0.02, *p*>0.05), polygenic overlap was substantial, with the majority of trait-influencing variants shared between the three traits. This contrast reflects mixed effect directions, which we show through convergent results from causal mixture modelling, local genetic correlations, and conjunctional false discovery rate analysis. Genes shared between activity and schizophrenia were differentially expressed in the amygdala and several basal ganglia structures, and associated with a range of cognitive-affective traits. In contrast, genes shared between activity and PD were largely associated with neurodegenerative disorders and structural brain phenotypes. Together, these findings suggest that activity draws on genes involved in motor, cognitive, affective and neurodegenerative processes, providing a genetic basis for its alteration across diverse neuropsychiatric disorders.

## 1. Introduction

Physical activity levels are altered across diverse neurological and psychiatric disorders [1–3]. Objectively measured activity is thus emerging as an informative transdiagnostic biomarker for outcomes including early diagnosis, disease progression, and functional status [1, 4–7]. The neurobehavioural processes underlying physical activity span motivation, arousal, and reward processing through to motor execution and control, many of which are disrupted in neuropsychopathology [8]. Consistent with a neural contribution, genome-wide association studies (GWAS) estimate that common variants account for ∼15% of population-level variance in physical activity, with associated loci enriched for genes expressed in the central nervous system [9–11]. Yet, few studies have investigated the shared genetic underpinnings of physical activity levels and neuropsychiatric disorders, which may yield insights into shared biological pathways that govern activity in both health and disease [12, 13].

Schizophrenia and Parkinson’s disease (PD) may be particularly informative comparisons for characterising this shared genetic architecture [5, 7, 14, 15]. Both disorders involve aberrant dopaminergic functioning within mesolimbic and nigrostriatal circuitry, which may reduce physical activity through overlapping mechanisms, including hypokinesia and amotivational syndromes [16–19]. These disturbances often correlate with clinically salient outcomes, such as negative symptoms in schizophrenia and quality of life in PD [2, 6, 7, 15]. Yet, the disorders are etiologically distinct. Schizophrenia is a highly polygenic neurodevelopmental disorder that affects synaptic function, whereas PD is substantially less polygenic and is primarily characterised by neurodegeneration of dopaminergic neurons in the substantia nigra [20–22]

Existing literature examining the genetic overlap between physical activity, schizophrenia, and PD is sparse and difficult to interpret [12, 13]. Polygenic risk scores for schizophrenia are associated with reduced overall activity, yet genetic correlation analyses point in the opposite direction, showing small positive correlations with time spent walking and negative correlations with sedentary behaviour [12]. Genetic correlations between activity and PD are negligible [13]. One possible explanation for these inconsistencies is that shared variants have a mixture of concordant and discordant effect directions, which cancel out to produce weak, null, or conflicting estimates in standard genetic correlation analyses [23]. Such a pattern was recently shown between PD and schizophrenia, where methods agnostic to effect directions estimated substantial polygenic overlap despite negligible genetic correlations [24, 25]. This consideration may be particularly relevant to schizophrenia, where motor dysfunction encompasses both hyperkinetic (e.g., psychomotor agitation, catatonic excitement) and hypokinetic (e.g., psychomotor slowing, stuporous catatonia) presentations, potentially implicating shared genetic variants with opposing effects on overall activity levels [26].

Here, we used GWAS summary statistics to characterise the genetic overlap among objectively measured physical activity, schizophrenia, and PD, using methods designed to detect shared genetic architecture irrespective of bulk genetic correlation [27–29]. We further sought to characterise the brain regions, biological pathways and associated traits implicated by specific shared loci [30]. To attain sufficient statistical power to apply these methods, we first enhanced existing accelerometer-derived activity GWAS by jointly analysing them with self-reported physical activity data [31, 32].

## 2. Methods and Materials

### 2.1 GWAS Summary Statistics

We used publicly available GWAS summary statistics for physical activity, schizophrenia, and PD, restricting analyses to European ancestry samples to match the available accelerometer-derived activity data (Supplementary Table 1).

#### Physical activity

Summary statistics for accelerometer-derived and self-reported physical activity came from GWAS in the UK Biobank (UKB), a prospective cohort of ∼500,000 participants aged 40–69 years [33]. The accelerometer GWAS used a UKB subsample who wore wrist-worn accelerometers continuously for 7 days [34]. Overall activity was derived as the mean acceleration vector magnitude across valid epochs, excluding non-wear time, a metric which has been validated against established measures of energy expenditure, and shows good long-term reproducibility [34–36]. The GWAS included 89,683 European-ancestry participants after quality control [10].

Three self-reported physical activity phenotypes were available from touchscreen questionnaires about frequency, intensity and duration of activity administered to the larger UKB cohort [11]: (1) Moderate to vigorous physical activity (MVPA), a continuous phenotype derived from the sum of total minutes per week of moderate physical activity multiplied by four, and vigorous physical activity multiplied by eight (corresponding to their respective metabolic equivalents); (2) Vigorous physical activity (VPA), a dichotomized phenotype where minutes per week of VPA were collapsed into those that reported more than 3 days of at least 25 minutes of VPA per week, and those that reported less than 3 days; (3) Strenuous Sports, a dichotomized measure, in which participants were collapsed into those that spend 2–3 days/week or more doing strenuous sports or other exercises, for a duration of 15–30 min or greater. After quality control, the sample sizes were: *n* = 377,234 for MVPA; *n* = 98,060 cases and 162,995 controls for VPA; *n* =124,842 cases and 225,650 controls for Strenuous Sports [11].

#### Schizophrenia

Summary statistics were obtained from the European-ancestry subsample of the Psychiatric Genomics Consortium wave 3 schizophrenia GWAS (53,386 cases, 77,258 controls) [21].

#### Parkinson’s disease

Summary statistics were obtained from the European-ancestry subsample of the Global Parkinson’s Genetics Program [37]. We used GWAS of both the overall sample (63,555 cases, 17,700 proxy cases, and 1,746,386 controls), which combines biobank and case-control studies, and a case-control-only subset (34,933 cases, 31,009 controls). The subset was used where sample overlap with UKB could inflate overlap estimates (see Section 2.6).

### 2.2 Multi-trait analysis of GWAS

We applied Multi-trait analysis of GWAS (MTAG) to boost the statistical power of our primary accelerometer-derived activity GWAS [32]. MTAG models the variance-covariance structure of effect estimates across traits, allowing correlated traits to share information and increase power. Sample overlap is accounted for by estimating the residual covariance from the intercepts of univariate and bivariate linkage disequilibrium score regression (LDSC). A genetic variance-covariance matrix is then estimated using a method of moments, conditional on this residual covariance. Bivariate linkage disequilibrium score regression (LDSC) is used to estimate these parameters. MTAG assumes all single-nucleotide polymorphisms (SNPs) share the same variance-covariance matrix across traits. While this assumption is rarely satisfied in practice, MTAG remains a consistent estimator with a lower genome-wide mean squared error than single-trait GWAS. However, effect estimates may be biased away from zero when SNPs have true effects on one trait but not another. We thus calculated a maximum false discovery rate (maxFDR) as an upper bound under potential assumption violations. Lead SNPs were identified following the FUMA protocol [30]. Independent significant SNPs were defined as those independent from one another at r²<0.6, and lead SNPs as the subset of these that were further independent at r²<0.1. Candidate SNPs, defined as those in linkage disequilibrium (r²≥0.6) with an independent significant SNP, were used to define the boundaries of each locus, and loci within 250 kb of one another were merged into a single locus.

### 2.3 SNP heritability and Genetic correlation

We used LDSC to estimate SNP heritability for the physical activity MTAG summary statistics and to calculate genetic correlations between activity, schizophrenia and PD [38].

### 2.4 Causal Mixture Modeling

We used the MiXeR toolset to characterize the polygenicity of physical activity, and its genetic overlap with schizophrenia and PD [28, 39, 40]*. Univariate MiXeR* models genetic architecture using point-normal mixture prior with “null” variants having no effect and “trait-influencing” variants having normally distributed effects [28, 31]. Maximum likelihood estimation yields parameters including polygenicity (proportion of trait-influencing variants), discoverability (variance of trait-influencing variant effect sizes), observed SNP heritability, and the number of trait-influencing variants explaining 90% of SNP heritability. *Trivariate MiXeR* extends this framework to three phenotypes, modeling SNP effects as a mixture of eight components: variants null for all three traits, variants influencing only one trait, variants shared by each pair of traits but not the third, and variants shared across all three traits, while accounting for sample overlap between GWAS [40]. Stability was assessed by examining the variation in polygenicity estimates across 16 independent runs, each using a different random subset of 300,000 SNPs. The median was taken as the point estimate, and the run with the smallest deviation from the median pattern was used to visualize the data as an Euler diagram, with ellipses scaled to the polygenicity of the corresponding phenotype. For each pair of traits, we further estimated the fraction of variants with concordant effects from the correlation of effect sizes within each shared component. An LD reference panel based on European individuals from 1000 Genomes Phase 3 was used.

### 2.5 Local analysis of covariate association

We used Local Analysis of [co]Variant Association (LAVA v0.1.5) for local genetic correlation analysis [29]. LAVA calculates local genetic correlation at semi-independent genomic regions defined by recombination boundaries (∼1-2.5 Mb in size). It first tests local SNP heritability in each region. Where regions are significantly heritable for both traits, bivariate genetic correlations are computed from local genetic covariances using a method of moments, while accounting for sample overlap. Statistical significance is assessed using simulation-based p-values. To correct for multiple testing across 2,495 regions, we applied a Bonferroni threshold of p<2×10^-5^. We used the UKB LD reference files provided for LAVA, and estimated sample overlap from LDSC intercepts.

### 2.6 Conjunctional False Discovery Rate

We used the conjunctional false discovery rate (conjFDR) approach to identify genomic loci jointly associated with each trait in the analysed pair of traits [41]. Cross-trait enrichment was assessed using conditional QQ plots, which visualise the expected versus observed p-values for a primary trait across SNP subsets stratified by their association strength with a secondary trait. The conjFDR statistic was computed for each genetic variant, defined as the posterior probability that a variant is not associated with one or both phenotypes, given that the p-values for both phenotypes are at least as small as their observed values. In the presence of cross-trait enrichment, this identified shared variants associated with both traits beyond genome-wide significance. Variants with conjFDR<0.05 were considered statistically significant. Lead SNPs and locus boundaries were identified following the FUMA protocol, as described in Section 2.2.

### 2.7 Functional annotation

Lead SNPs were mapped to genes using the Variant-to-Gene (v2g) tool from Open Targets Genetics [42]. V2g prioritises variants and identifies potential causal genes by integrating multiple types of evidence, including positional information, chromatin interaction data, expression, protein, and splicing quantitative trait loci, and in silico functional predictions, into a single composite score. The highest-scoring gene was assigned to each lead SNP. FUMA’s Gene2Func function was then used for gene-set enrichment analyses via hypergeometric tests [30]. Genes were investigated for enrichment in the Molecular Signatures database gene sets [43], comprising Gene Ontology terms (biological processes, molecular functions, and cellular components) and curated canonical pathways (KEGG and Reactome). Additionally, the genes were tested for enrichment in differentially expressed gene sets from the Genotype-Tissue Expression (GTEx) version 8 [44] and in genes previously reported in the GWAS Catalog as associated with other traits [45]. Multiple testing correction was applied within each gene set category using the Benjamini–Hochberg procedure.

### 2.8 Ethical Considerations

This study analysed only publicly available, de-identified GWAS summary-level data. The present secondary analyses were approved by the University of Cape Town Faculty of Health Sciences Human Research Ethics Committee (HREC ref. 012/2025).

## 3. Results

### 3.1 Multi-Trait Analysis of GWAS boosts the power of physical activity GWAS

The accelerometer-derived activity trait was moderately correlated with each self-reported trait (r_g_=0.41-0.5, all p<0.001; Supplementary Table 2), supporting the use of MTAG to boost discovery using self-report GWAS. The resulting physical activity MTAG showed an increased effective sample size (*n*=107,489 versus 89,683) and mean chi-squared (1.25 versus 1.21; Supplementary Table 3) relative to accelerometer-derived activity alone. A maxFDR value of 0.01 supported the reliability of these results. Clumping produced 15 lead SNPs (of 27 independent significant SNPs) in 14 genome-wide significant loci, versus 9 independent significant SNPs in the original GWAS [10] (Figure 1a; Supplementary Table 4). Lead SNPs mapped to 14 genes, based on the top v2g score for each SNP using Open Targets (Supplementary Table 5).

**Figure 1:**
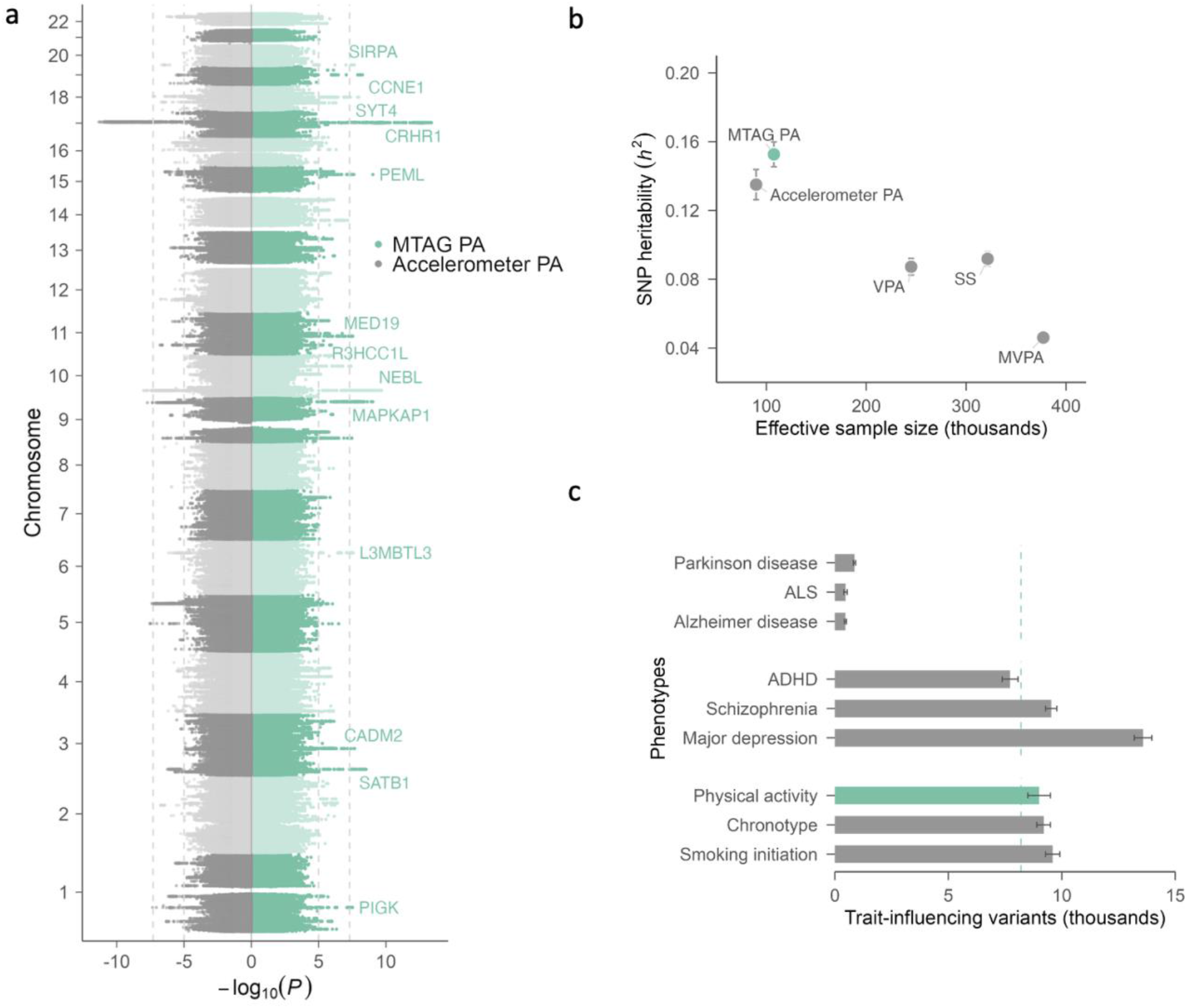
The Genetic Architecture of Physical Activity (a) Miami plot comparing MTAG physical activity GWAS (green) with accelerometer-derived physical activity alone (grey), demonstrating the power boost from MTAG. The *y*-axis indicates chromosomal position; the *x*-axis shows −log₁₀(*p*). Dashed lines mark the suggestive (1×10^-5^) and genome-wide significance (5×10^-8^) thresholds. Lead SNPs are annotated with associated genes based on the top Open Targets v2g score. (b) SNP heritability (*h*²) plotted against effective sample size. MTAG PA (green) achieves a boost in sample size over accelerometer PA while preserving high heritability, in contrast to self-reported traits (VPA, SS, MVPA; grey), which have larger samples but lower heritability. (c) Bar plot comparing the polygenicity of the PA GWAS (turquoise) to polygenicity estimates, provided as the number of trait-influencing variants explaining 90% SNP-based heritability of selected psychiatric, behavioral and neurological traits adapted from published reports (grey). Error bars show standard deviations across multiple independent runs. PA, physical activity. VPA, vigorous physical activity. SS, strenuous sports. MVPA, moderate to vigorous physical activity. ALS, amyotrophic lateral sclerosis. ADHD, attention deficit hyperactivity disorder.

### 3.2 Physical activity is moderately heritable and highly polygenic

The physical activity MTAG had a SNP heritability of 0.15 (SE=0.0072) as per LDSC (Figure 1b). Univariate MiXeR indicated that the trait was highly polygenic, with ∼9K (SD: 0.5K) trait-influencing variants estimated to account for 90% of the SNP heritability (Figure 1c, Supplementary Table 6). At the present sample size, genome-wide significant SNPs explain approximately 0.3% of the SNP heritability of physical activity (Supplementary Figure 1). Robustness checks supported the suitability of the MTAG-boosted summary statistics for univariate MiXeR analysis over the non-boosted version (Supplementary Table 6; Supplementary Figure 2).

### 3.3 Trivariate MiXeR revealed extensive polygenic overlap

Genetic correlations between physical activity and both schizophrenia and PD traits were negligible (r_g_=-0.01–0.02, *p*>0.05; Supplementary Table 7). Nevertheless, the trivariate MiXeR model revealed extensive shared genetic architecture (Figure 2). Of the total variants influencing the three traits, 9% were shared across all three, and 56% between schizophrenia and physical activity. Variants shared between PD and activity were entirely subsumed within the schizophrenia overlap. Less than 1% of variants were unique to PD, and only 12% and 29% of variants were unique to physical activity and schizophrenia, respectively. The proportion of shared variants with concordant effect directions was 50% between physical activity and PD, 53% between physical activity and schizophrenia, and 54% between schizophrenia and PD. Across 16 runs, estimates of the SCZ-specific component and of the PA–SCZ, PD–SCZ and three-way overlaps were stable (relative half interquartile range [RHIQR] 0.17–0.47). In contrast, the smaller PA-specific and PD-specific components and the PA–PD overlap showed greater run-to-run variability (RHIQR 0.70 1.88) and should be interpreted with caution (Supplementary Table 9).

**Figure 2:**
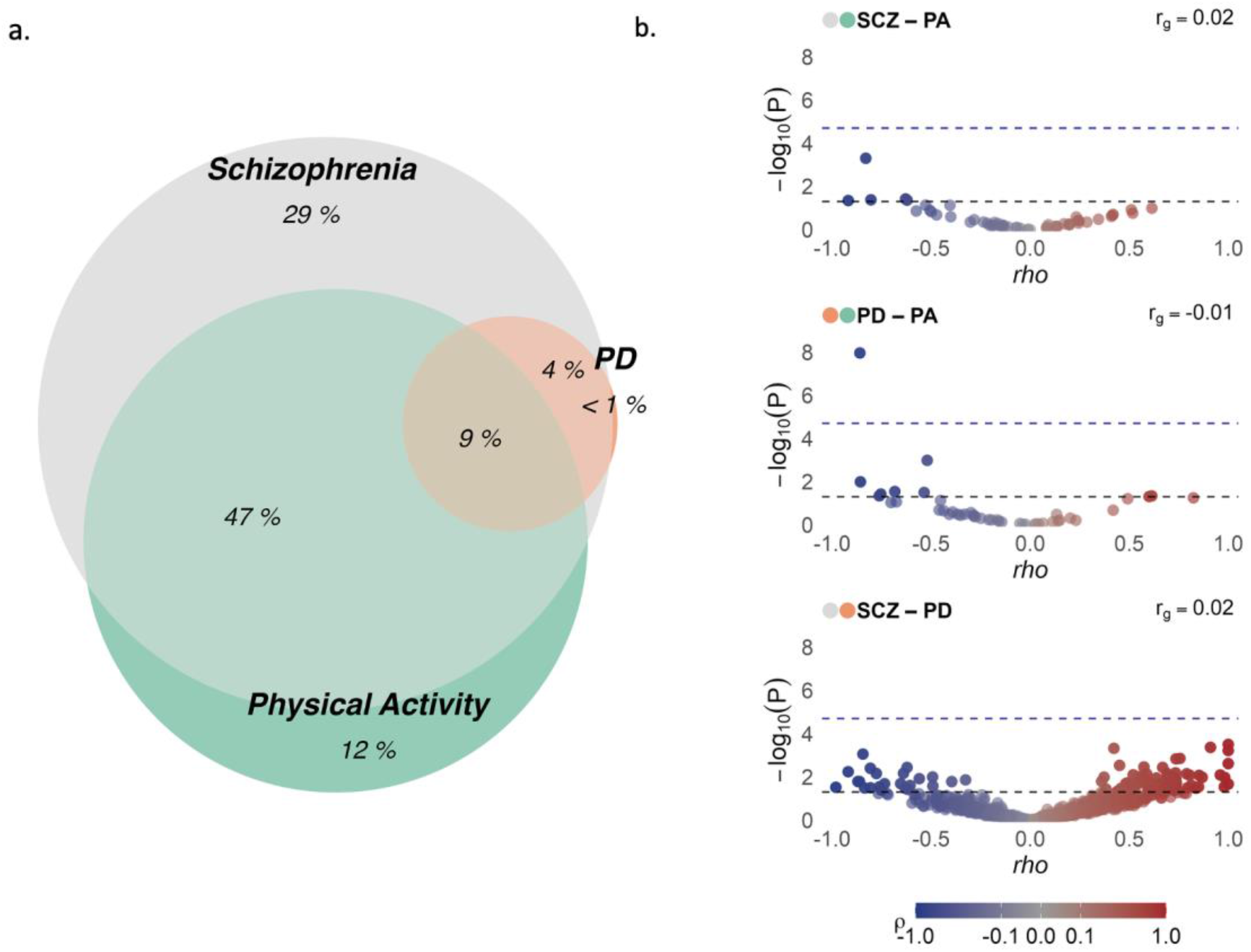
Genome-wide genetic overlap between physical activity, schizophrenia and Parkinson’s disease (a) Venn diagram showing trivariate MiXeR-modeled genome-wide genetic overlap between physical activity (green), schizophrenia (grey) and Parkinson’s disease (orange). Percentages represent each region’s proportion of the total trait-influencing variants estimated across the three phenotypes. (b) Volcano plots showing LAVA local genetic correlation results. Each point represents a genomic region; the *x*-axis shows local genetic correlation (*ρ*) and the *y*-axis shows significance (−log₁₀(*p*)). Points are coloured by the direction and magnitude of the correlation (blue = negative, red = positive). Dashed lines indicate the nominal significance level (0.05) and Bonferroni-corrected significance level (2×10^-5^). Each plot is labelled with its trait pair, with the two coloured dots matching the phenotype colours in panel (a); the genome-wide genetic correlation (*r_g_*) for each pair is shown at top right. SCZ, schizophrenia. PA, physical activity. PD, Parkinson’s disease.

### 3.4 Bivariate LAVA implicated the 17q21.31 region as a pleiotropic hotspot

Among genomic regions with significant univariate heritability for both traits, 33% (15/45) showed positive local genetic correlations between physical activity and PD, 44% (25/57) between physical activity and schizophrenia, and 61% (525/924) between schizophrenia and PD (Figure 2b-d). In the PD-activity analysis, one region on chromosome 17 (positions 43460501–44865831) surpassed the Bonferroni threshold (rho=-0.86; *p*=1.1×10^-8^) (Figure 3a). The same region was nominally significant in the schizophrenia–activity (rho=-0.83; *p*=5×10^-4^) and schizophrenia–PD analyses (rho=0.42; *p*=6.01×10^-4^). Nominally significant genetic regions across each trait pair are reported in Supplementary Tables 10–12.

**Figure 3:**
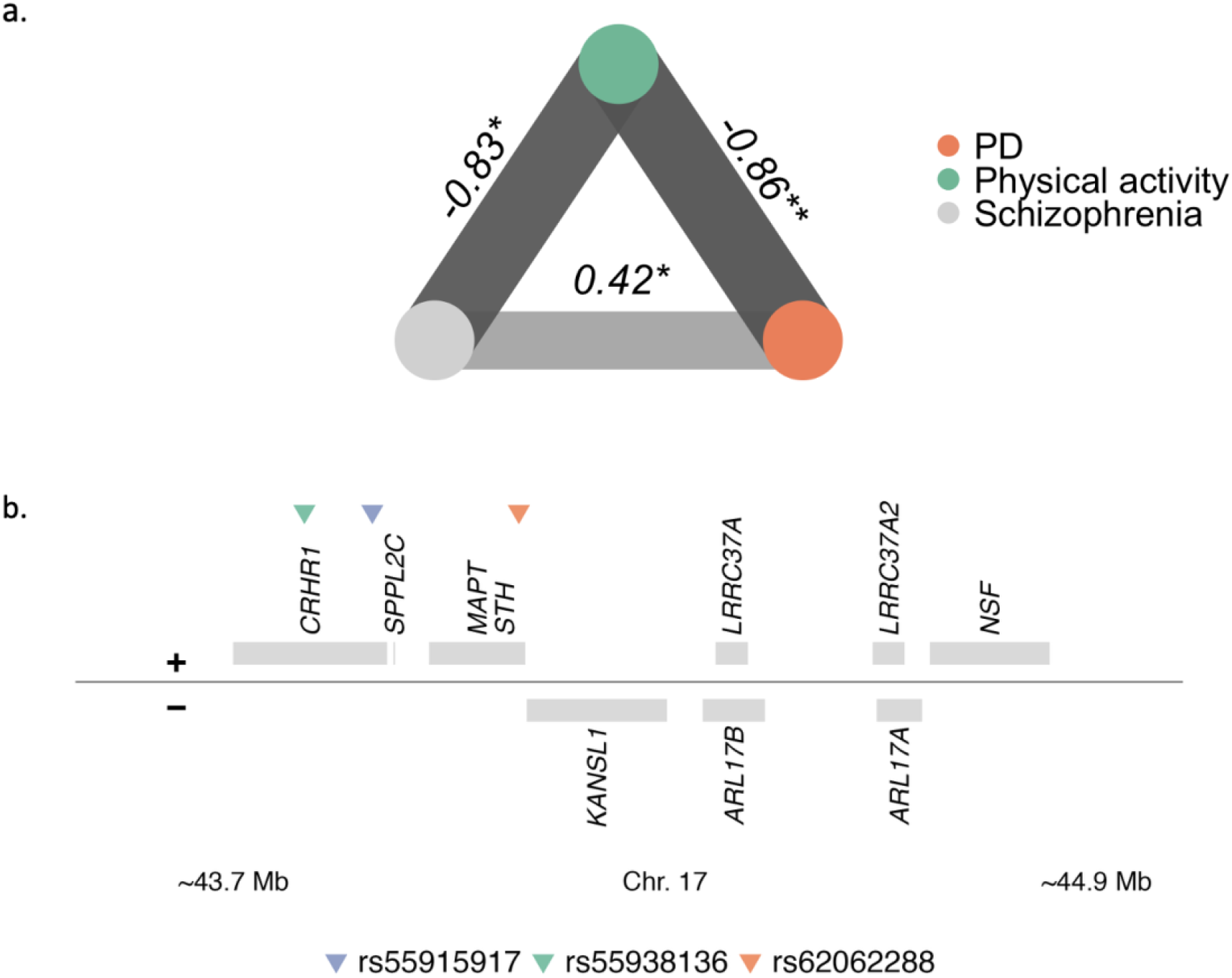
Convergence of signal at 17q21.31 (a) Network plot of local genetic correlations at the 17q21.31 locus. Edge values show local genetic correlations; edge width and opacity reflect association strength. \**p*<0.05; \*\**p*<2×10^-5^. (b) Gene map of a portion of the 17q21.31 locus (∼43.7–44.9 Mb) with protein-coding genes on forward (+) and reverse (-) strands. Triangles indicate conjFDR-significant SNPs in this region. PD, physical activity. Chr., chromosome.

### 3.5 Conjunctional FDR identified specific shared variants with physical activity

ConjFDR identified 32 lead SNPs in 31 loci shared between schizophrenia and physical activity (FDR < 0.05), of which 38% had concordant effect directions, and 11 lead SNPs in 10 loci shared between PD and physical activity, of which 64% had concordant effect directions (Supplementary Tables 13 and 14). Genes mapped to each SNP by Open Targets top v2g score are listed in Supplementary Tables S15 and S16. Variant rs55938136, which maps to the *MAPT* gene, showed pleiotropy across both trait pairs. This variant falls within the chromosome 17 region implicated in the LAVA analyses (positions 43,460,501-44,865,831). Other independent pleiotropic SNPs within the same locus included rs62062288, mapping to *KANSL1* (schizophrenia and physical activity), and rs55915917, mapping to *CRHR1* (PD and physical activity).

### 3.6 Gene-set enrichment of pleiotropic genes

#### Differential tissue expression

Genes overlapping between schizophrenia and physical activity showed significant differential expression between brain and non-brain tissues, most strongly in the amygdala and several basal ganglia structures (putamen, nucleus accumbens, caudate, and substantia nigra), with additional enrichments in the hippocampus and anterior cingulate cortex (Supplementary Figure 3). In contrast, genes overlapping between PD and physical activity did not show any significant differential expression across tissue types (Supplementary Figure 4).

#### Molecular signatures database

A single gene ontology term, “neuron projection,” was enriched in the PD pleiotropic gene set (adjusted *p*=0.01), driven by 6 of 11 shared genes (*CRIPT*, *PHLPP2, CRHR1, MAPT, ACTG1,* and *RIT2)*. No other gene sets surpassed the significance threshold.

#### GWAS catalogue

Enrichment for previously associated traits revealed both shared and gene-set-specific signals (Figure 4; Supplementary Figures 5–6). Both gene sets were enriched for substance use and neuroticism. The schizophrenia–activity set was additionally enriched for mental health, cognitive, and anthropometric traits (including BMI), and the PD–activity set for neurological disorders and structural brain phenotypes. Nearly all PD–activity enrichments were driven by *CRHR1* and *MAPT*, both within the 17q21.31 region.

**Figure 4:**
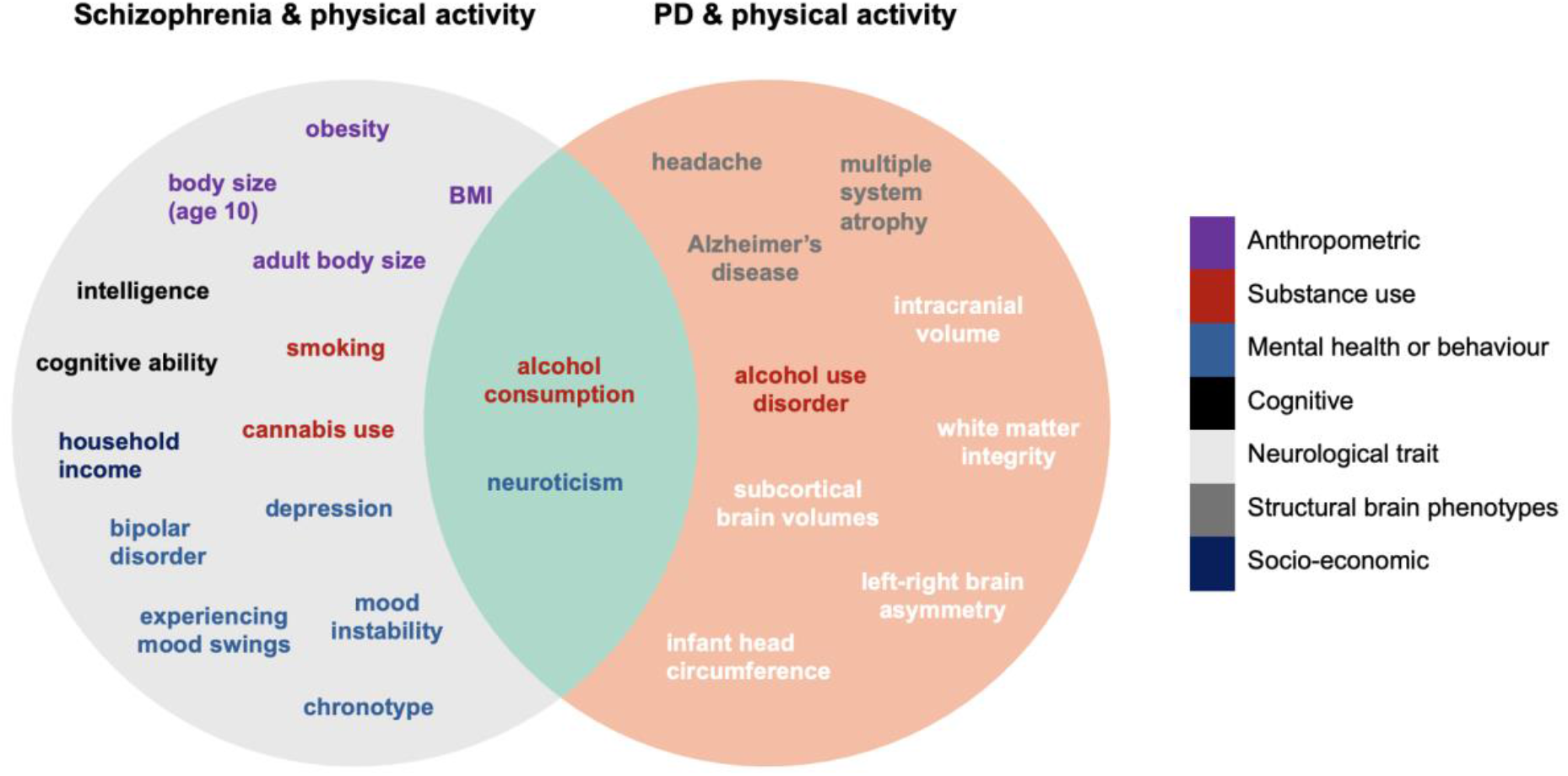
Venn diagram of GWAS catalogue trait enrichments for genes shared between schizophrenia and physical activity (left) and between Parkinson’s disease and physical activity (right). The overlap (green) shows traits enriched in both gene sets. Traits are colour-coded by category (legend). Full results in Supplementary Figures 6–7.

## 4. Discussion

Physical activity levels are altered across neuropsychiatric disorders, yet the genetic basis for this link is poorly characterised. Here, we found that physical activity is highly polygenic and draws from a largely shared pool of common variants with schizophrenia and PD. These shared variants had mixed effect directions that cancel out in aggregate, yielding negligible genetic correlations. Resolving the overlap to individual genomic regions identified a pleiotropic hotspot within 17q21.31. Gene set analyses of shared genes implicated brain regions involved in both motor and cognitive-affective function, and associated traits across structural brain phenotypes, neurological and psychiatric disorders, cognition, and behaviour.

Joint analysis of the accelerometer-derived physical activity GWAS and the self-reported activity GWAS using MTAG boosted the power of the original GWAS while preserving its higher heritability relative to self-reported data. This extends previous work using MTAG to boost power for more deeply phenotyped samples via scalable proxies, here applied to a wearable-derived phenotype [32]. The resulting increase in power enabled us to characterise the genetic overlap of physical activity with schizophrenia and PD using tools such as MiXeR [31].

Physical activity is highly polygenic, with univariate MiXeR estimating 9000 trait-influencing variants, on the order of other complex behavioural traits such as smoking initiation and mental disorders such as schizophrenia (both ∼9,600 variants) [23, 46]. In contrast, PD involves a tenfold lower polygenicity (∼900 variants), comparable to other neurodegenerative disorders such as Alzheimer’s disease (∼400 variants) (Figure 1c) [25]. Across all three traits, few variants were trait-specific. This was most pronounced for PD, though this should be interpreted in the context of its lower polygenicity. Nearly all variants influencing PD were shared with schizophrenia (<1% unique to PD), as was the entire PD-physical activity overlap. Likewise, schizophrenia- and physical-activity-specific variants each made up a small share of the total variation across the three traits (29% and 12%, respectively). Critically, this overlap was characterised by mixed effect directions, accounting for the negligible genetic correlations between these traits.

These results add to a growing body of evidence of genetic overlap extending beyond genetic correlation among complex psychiatric, neurological, and behavioural traits [23, 25, 47]. Rather than being distinguished by trait-unique variants, these traits may differ in the particular configuration of effect directions and magnitudes across a largely shared pool of variants [23]. To the extent that these variants do converge on shared biological pathways, complex neuropsychiatric phenotypes may be better understood as aggregates of shared mechanisms than as discrete entities, consistent with transdiagnostic frameworks such as the Research Domain Criteria [23, 48]. Physical activity may represent a behavioural dimension on which multiple shared mechanisms converge [49, 50].

At finer resolution, one shared locus showed significant discordant local genetic correlations between activity and PD, with nominal correlations between schizophrenia and both PD and activity. The locus spans the well-characterised 17q21.31 inversion polymorphism, a ∼900kb segment exhibiting extended linkage disequilibrium across multiple genes [51]. ConjFDR analysis identified multiple pleiotropic variants within this region. While the region’s unusual LD structure complicates causal attribution to a single gene, it is notable that Open Targets prioritised the genes *CRHR1*, *MAPT*, and *KANSL1*, which have all been previously linked to neurodevelopmental or neurodegenerative phenotypes [52–54]. These findings highlight the 17q21.31 region as a potential point of convergence between reduced physical activity and neuropsychiatric traits.

Variants shared with schizophrenia mapped to genes differentially expressed in the amygdala and several basal ganglia structures. The convergence of schizophrenia and physical activity loci in these regions may relate to the roles of the basal ganglia and amygdala in linking motivational and affective processing, respectively, to motor output. The basal ganglia integrate motor, cognitive, and motivational control through parallel cortico-striatal loops and, notably, pharmacological perturbation of nigrostriatal circuitry produces motor symptoms in schizophrenia [55–59]. The amygdala, in turn, drives motor behaviour in response to emotional stimuli, as in the ‘fight, flight, or freeze’ response [60, 61]. Abnormal motor behaviours across psychiatric disorders have been conceptualised as aberrant or extreme manifestations of these responses under severe psychological distress [62, 63].

Genes shared between physical activity and both disorders converged on enrichment for substance use and neuroticism. Such enrichments could plausibly arise through at least two routes. They may reflect shared upstream mechanisms. For example, reward pathways may independently influence substance use and normal variation in physical activity [64]. This aligns with the conceptualisation of aberrant activity alongside comorbid substance use, negative symptoms in schizophrenia, and the related symptoms of amotivation and anhedonia in PD as symptomatic manifestations of dopaminergic dysfunction [17, 65–67]. Alternatively, they may reflect causal links between the phenotypes themselves. Mood disturbances, which are common in both schizophrenia and PD, may reduce activity, while reduced activity may in turn worsen mood [68–70]. Both routes likely contribute to these enrichments.

Relatedly, genes shared between schizophrenia and physical activity were enriched for multiple BMI-related traits, with nearly all underlying genes (*ERBB4, CADM2, MEF2C, DOCK1, NCAM1, PCDH17*) showing concordant effect directions. Genes enriched for associations with BMI were linked to increased schizophrenia risk and increased activity. This is notable given the paradoxically negative genetic correlations previously reported between schizophrenia and BMI, despite substantial clinical comorbidity, which is thought to reflect the impact of lifestyle factors over genetic factors in driving this comorbidity [71].

Gene-set analyses also revealed contrasting enrichments between schizophrenia- and PD-shared genes. Variants shared between activity and schizophrenia were generally enriched for traits related to cognition, mental health, and behaviour (e.g., cognitive ability, bipolar disorder, chronotype). In contrast, variants shared between activity and PD were primarily enriched for genes associated with neurological disorders and structural brain phenotypes (e.g., Alzheimer’s Disease, multiple system atrophy, white matter integrity, subcortical brain volumes). Taken together, these shared and distinct enrichments are consistent with the psychomotor nature of physical activity, a phenotype shaped by motivational, cognitive, and affective influences as well as the structural substrates of movement.

### Limitations

This study has some limitations. First, the GWAS summary statistics were derived from samples of European ancestry, limiting generalisability to other populations. Second, trivariate MiXeR estimates involving Parkinson’s disease showed substantial run-to-run variability and should therefore be interpreted with caution. Finally, tissue-level gene-set enrichment analyses capture aggregate patterns that do not account for cell-type specificity, cell state or developmental timing. Inferences regarding shared biological mechanisms therefore remain hypothesis-generating.

### Conclusions

Physical activity is highly polygenic and shares substantial genetic architecture with schizophrenia and PD, despite negligible genetic correlations. This overlap is characterised by mixed effect directions and gene-set enrichments spanning motor, cognitive, affective and neurological phenotypes. Together, these findings highlight the complex genetic architecture underlying physical activity and suggest that it reflects the convergence of multiple neurobehavioural processes also implicated in neuropsychiatric disorders.

## Data Availability

All data produced in the present study are available upon reasonable request to the authors.

## 5. Acknowledgements

The work reported herein was made possible through funding by the South African Medical Research Council (SAMRC) through its Division of Research Capacity Development under the SAMRC Clinician Researcher Development Programme, with funding received from the National Department of Health. We thank Associate Professor Henk Temmingh for his feedback on later drafts and stimulating discussions on the clinical interpretation of these findings. We dedicate this work to the memory of Professor Dan Stein, whose mentorship was foundational to this project. The authors used Claude Opus 4.6 (Anthropic) to provide suggestions to improve the flow and readability of the text. All suggestions were critically reviewed by the authors, who take full responsibility for the scientific content and final manuscript.

