## Supplementary Figures for "Polygenic overlap of physical activity with schizophrenia and Parkinson’s disease"

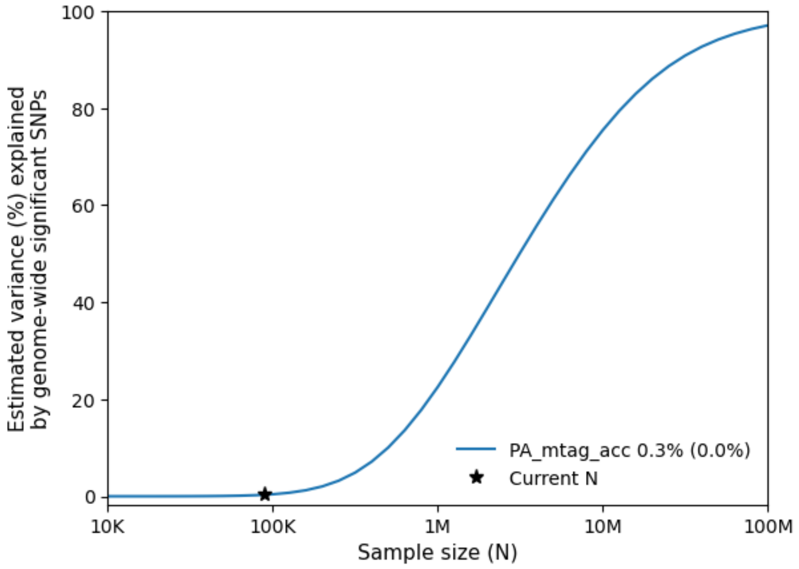


Supplementary Figure 1: Power curve for the physical activity MTAG (PA_mtag_acc) estimated using univariate MiXeR. The y-axis shows the proportion of SNP-heritability explained by genome-wide significant SNPs ($p < 5{\times}10^{-8}$) as a function of GWAS sample size (x-axis, log scale). The blue curve represents the MiXeR prediction, with the star indicating the current sample size. At the present sample size, genome-wide significant SNPs explain approximately 0.3% of SNP-heritability. The gradual slope of the curve indicates low discoverability, such that very large samples would be required to capture a substantial proportion of genetic variance.

A

B

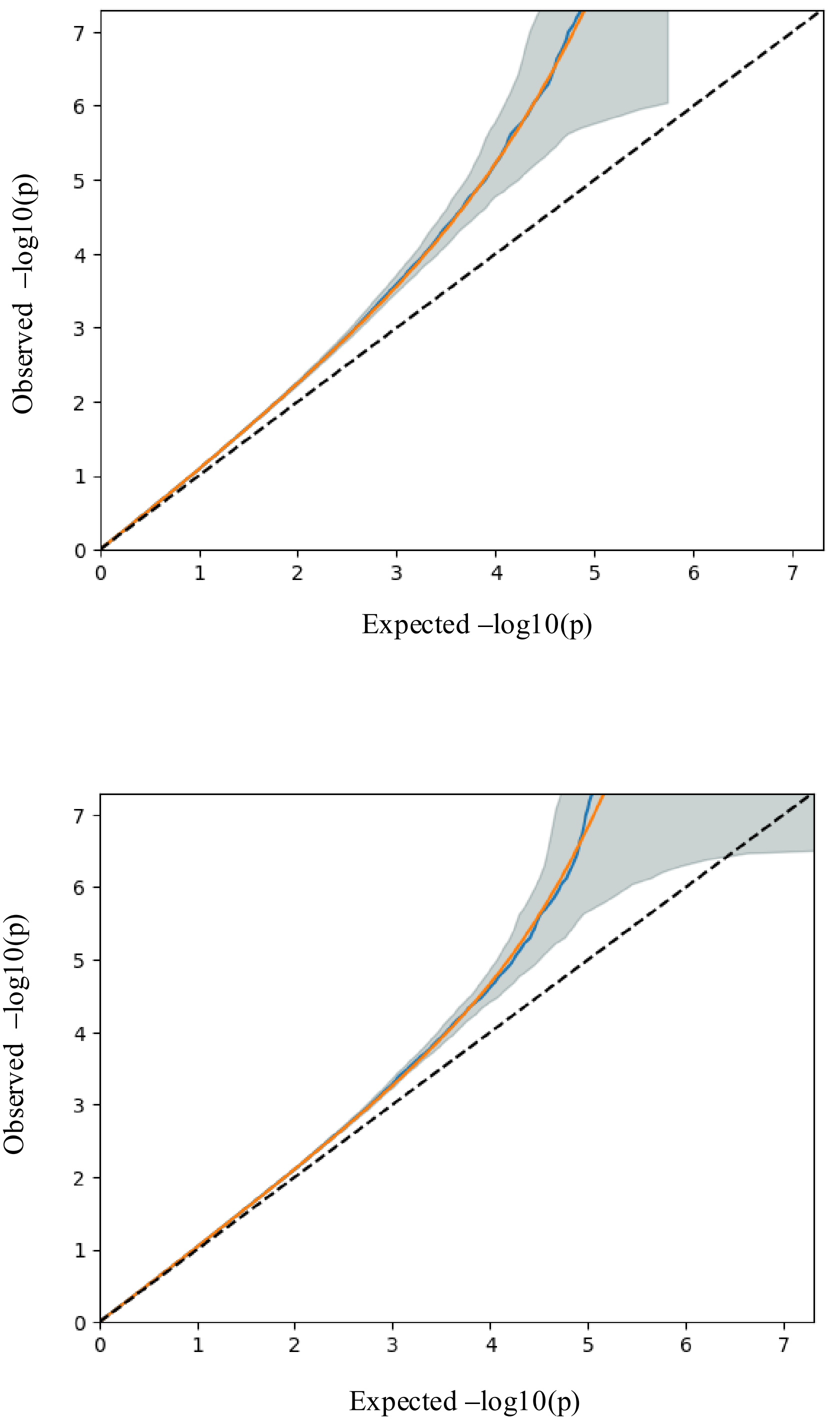


Supplementary Figure 2: QQ plots describing univariate MiXeR model fit for the physical activity MTAG. Panel A: *Fit plot*, based on a subset of linkage disequilibrium-pruned SNPs used to estimate model parameters. Panel B: *Test plot*, based on the entire set of SNPs. The blue line shows the observed GWAS distribution, the yellow line shows the predicted distribution from the fitted MiXeR model, the grey shaded area represents the 95% prediction interval, and the dashed diagonal represents the null hypothesis of no association. The departure of the GWAS curve from the null line reflects SNP-heritability; its curvature indicates polygenicity (the point where the curve bends away from the null), and the slope after departure reflects discoverability. Consistency between fit and test plots indicates that the model generalizes beyond the training data.

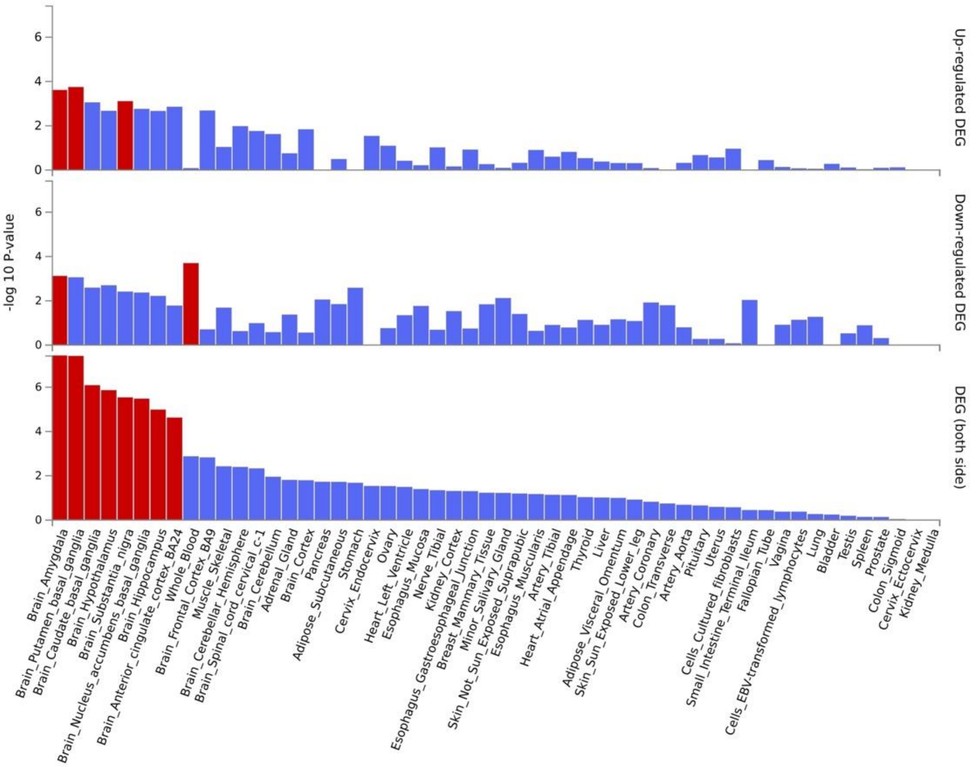


Supplementary Figure 3: Gene set enrichment analysis of pleiotropic genes shared between schizophrenia and physical activity for differential gene expression in GTEx v8 54 tissue types

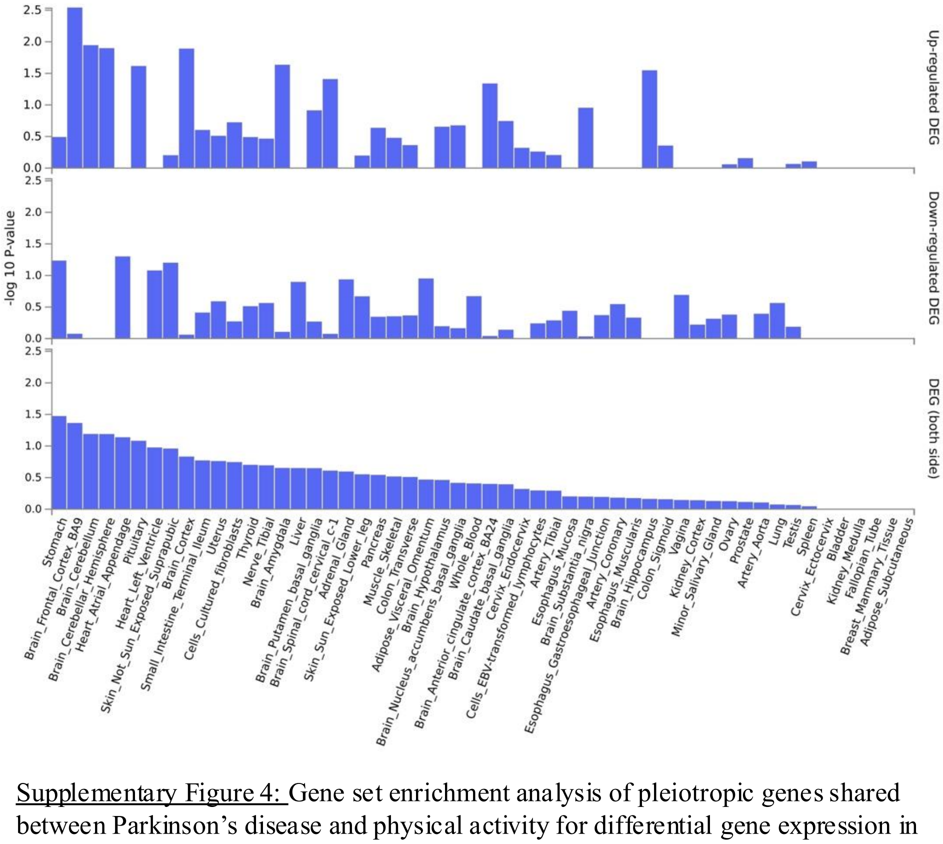


Supplementary Figure 4: Gene set enrichment analysis of pleiotropic genes shared between Parkinson’s disease and physical activity for differential gene expression in GTEx v8 54 tissue types

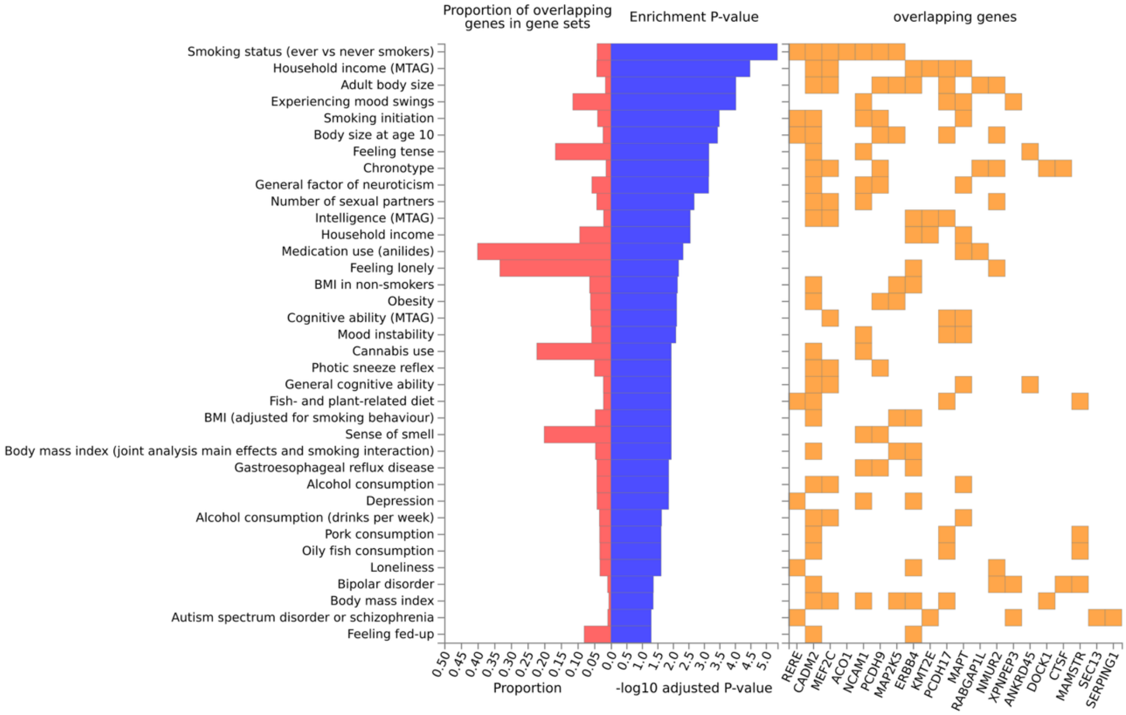


Supplementary Figure 5: Gene set enrichment analysis of pleiotropic genes shared between schizophrenia and physical activity for previously associated genes in the GWAS catalogue

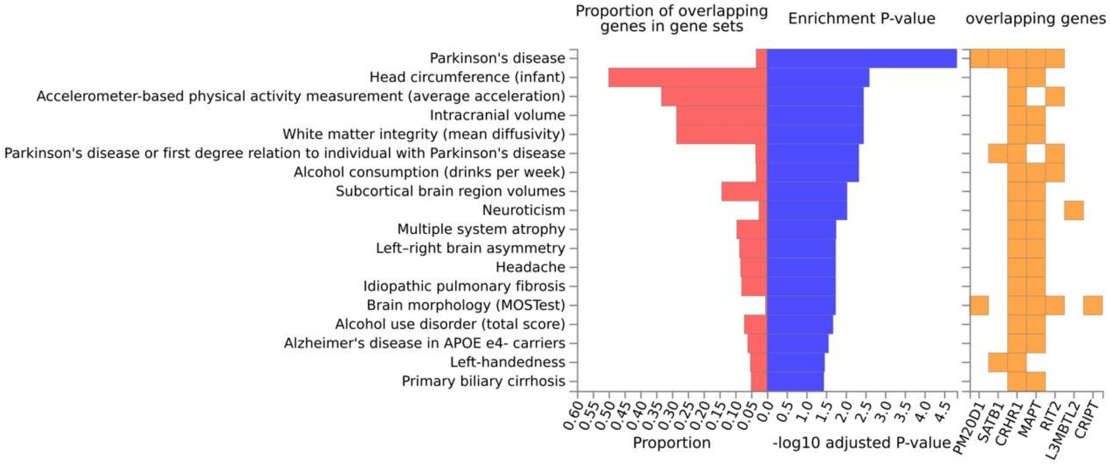


Supplementary Figure 6: Gene set enrichment analysis of pleiotropic genes shared between Parkinson's disease and physical activity for previously associated genes in the GWAS catalogue
